# Epigenetic profiling of Italian patients identified methylation sites associated with hereditary Transthyretin amyloidosis

**DOI:** 10.1101/2020.04.13.20064006

**Authors:** Antonella De Lillo, Gita A. Pathak, Flavio De Angelis, Marco Di Girolamo, Marco Luigetti, Mario Sabatelli, Federico Perfetto, Sabrina Frusconi, Dario Manfellotto, Maria Fuciarelli, Renato Polimanti

## Abstract

Hereditary Transthyretin (TTR) Amyloidosis (hATTR) is a rare life-threatening disorder caused by amyloidogenic coding mutations located in *TTR* gene. To understand the high phenotypic variability observed among carriers of *TTR* disease-causing mutations, we conducted an epigenome-wide association study (EWAS) assessing more than 700,000 methylation sites and testing epigenetic difference of *TTR* coding mutation carriers *vs*. non-carriers, We observed a significant methylation change at cg09097335 site located in *Beta-secretase 2* (*BACE2*) gene (beta =-0.60, p=6.26×10^−8^). This gene is involved in a protein interaction network enriched for biological processes and molecular pathways related to amyloid-beta metabolism (Gene Ontology:0050435, q=0.007), amyloid fiber formation (Reactome HSA-977225, q=0.008), and Alzheimer’s disease (KEGG hsa05010, q=2.2×10^−4^). Additionally, *TTR* and *BACE2* share APP (Amyloid-beta precursor protein) as a validated protein interactor. Within *TTR* gene region, we observed that Val30Met disrupts a methylation site, cg13139646, causing a drastic hypomethylation in carriers of this amyloidogenic mutation (beta=-2.18, p=3.34×10^−11^). Cg13139646 showed co-methylation with cg19203115 (r^2^=0.32), which showed significant epigenetic differences between symptomatic and asymptomatic carriers of amyloidogenic mutations (beta=-0.56, p=8.6×10^−4^). In conclusion, we provide novel insights related to the molecular mechanisms involved in the complex heterogeneity of hATTR, highlighting the role of epigenetic regulation in this rare disorder.

## Background

Hereditary transthyretin amyloidosis (hATTR; OMIM#105210) is a life-threatening disorder caused by transthyretin (TTR) misfolding and consequently amyloid fibril deposition in several tissues (e.g., peripheral nerves, heart, and gastrointestinal tract) (1, 2). This rare condition is characterized by extreme clinical heterogeneity including age of onset, penetrance, and clinical display (3-5). To date, more than 130 amyloidogenic mutations have been identified in the coding regions of the *TTR* gene, which are the cause of hATTR (6). The prevalence of hATTR is estimated to be approximately 1/100,000 (7). However, endemic areas of hATTR were identified in Portugal and Sweden (4, 5). Although both of these regions are affected by the same amyloidogenic mutation, Val30Met (rs28933979), the penetrance and age of onset are different: early age of onset and high penetrance in Portugal (4, 5, 8, 9) *vs*. late age of onset and low penetrance in Sweden and in non-endemic countries (3, 10, 11). It has been hypothesized that hATTR phenotypic heterogeneity is due to the contribution of genetic and non-genetic factors involved in the complex genotype-phenotype correlation observed (12-18). Recent data strongly support the role of non-coding regulatory variation on *TTR* gene expression, as one of the mechanisms affecting the phenotypic manifestations observed in carriers of *TTR* amyloidogenic mutations (19-22). Among genomic regulatory features, epigenetic modifications are demonstrated to be key mechanisms in modulating a wide range of molecular functions and potential targets to develop novel treatments (23-25). Of several epigenetic modifications, DNA methylation is the most studied with respect to human traits and diseases (23). With respect to monogenic disorders, methylation studies investigate the role of epigenetic changes involved in the phenotypic expression observed among carriers of disease-causing mutations (26-28). While epigenetic modifications have the potential to be involved in hATTR pathogenic mechanisms, to our knowledge no study has explored methylation changes of patients affected by this life-threatening disease. In the present study, we conducted an epigenome-wide association study (EWAS) to identify DNA methylation sites associated with hATTR, investigating 48 carriers of *TTR* amyloidogenic mutations and 32 controls. We also tested whether there are significant epigenetic changes among carriers of different amyloidogenic mutations. The results obtained showed: i) hATTR confirmed cases have significant methylation changes in modifier genes potentially involved in amyloidogenic processes; ii) carriers of Val30Met mutation showed a significant hypomethylation in *TTR* gene when compared to the carriers of other *TTR* amyloidogenic mutations.

## Results

The epigenome-wide analysis testing the differences between 48 carriers of *TTR* amyloidogenic mutations and 32 controls identified a significant methylation site surviving false discovery rate multiple testing correction (FDR q<0.05) at the cg09097335 site located in *Beta-secretase 2* (*BACE2*) gene body (beta = −0.60, *p*=6.26×10^−8^, FDR q=0.044). Carriers of *TTR* amyloidogenic mutations showed a significant hypomethylation when compared to controls (Figure 1). To understand whether methylation at this CpG site is associated with disease-associated genetic differences or post-disease processes, we compared hATTR patients, asymptomatic carriers of *TTR* mutations, and controls. Significant differences were observed for i) hATTR patients *vs*. controls (beta=-0.402, *p*=5.7×10^−4^) and ii) asymptomatic carriers vs. controls (beta=-0.716, *p*=3.21×10^−5^), but no difference was present between hATTR patients *vs*. asymptomatic carriers (beta=0.137, *p*=0.332). Leveraging GTEx data (29), we observed a complementary transcriptomic regulation between *TTR* and *BACE2* genes where the first is mainly expressed in its source organ (i.e. liver) while the second is expressed in target organs of TTR amyloid deposits (Figure 2). We investigated interactive proteins related to *TTR* and *BACE2* loci based on multiple experimental and computational evidence, identifying five candidates with medium-to-highest interaction confidence (Figure 2B). These include FYN (FYN proto-oncogene, Src family tyrosine kinase; interaction score=0.809), BACE1 (Beta-secretase 1; interaction score=0.804), APP (amyloid-beta precursor protein; interaction score=0.430), IGHV3-11 (immunoglobulin heavy variable 3-11; interaction score=0.412), and ENSG00000259680 (uncharacterized protein similar to an immunoglobulin heavy variable 3/OR16 gene; interaction score=0.412). Among them, TTR showed the highest interaction with APP protein (interaction score=0.936). BACE2 protein interactive network (Figure 3) showed functional enrichments for several biological processes and molecular pathways (Table 1). Among the enrichments directly related to BACE2 function that survived false discovery rate (FDR) multiple testing correction, we observed: Alzheimer’s disease (KEGG hsa05010, FDR q=2.2×10^−4^) related to the interaction of BACE2 with APP and BACE1; membrane protein ectodomain proteolysis (GO:0006509, FDR q=0.007) and amyloid-beta metabolic process (GO:0050435, FDR q=0.007) related to BACE2-BACE1 interaction; protein metabolic process (GO:0019538, FDR q=0.043) related to the interaction of BACE with APP, BACE1, FYN, and IGHV3-11. The interactions of other proteins within BACE2 interactive network also highlighted amyloid-related functional enrichments: amyloid fiber formation (Reactome HSA-977225, FDR q=0.008) and response to amyloid-beta (GO:1904645, FDR q=0.009) related to the interaction of APP with BACE1 and FYN, respectively.

**Table 1:** Enrichments for gene ontologies (GO) of biological processes and for Reactome and KEGG molecular pathways (HSA and hsa, respectively).

| ID | Description | Proteins | False Discovery Rate |
| --- | --- | --- | --- |
| hsa05010 | Alzheimer's disease | APP,BACE1,BACE2 | 2.2E-04 |
| HSA-2029481 | FCGR activation | FYN,IGHV3-11 | 7.9E-04 |
| HSA-2730905 | Role of LAT2/NTAL/LAB on calcium mobilization | FYN,IGHV3-11 | 7.9E-04 |
| HSA-983695 | Antigen activates B Cell Receptor (BCR) leading to generation of second messengers | FYN,IGHV3-11 | 0.002 |
| GO:0006509 | membrane protein ectodomain proteolysis | BACE1,BACE2 | 0.007 |
| GO:0050435 | amyloid-beta metabolic process | BACE1,BACE2 | 0.007 |
| GO:1902950 | regulation of dendritic spine maintenance | APP,FYN | 0.007 |
| HSA-977225 | Amyloid fiber formation | APP,BACE1 | 0.008 |
| GO:1904645 | response to amyloid-beta | APP,FYN | 0.009 |
| HSA-109582 | Hemostasis | APP,FYN,IGHV3-11 | 0.010 |
| GO:0106027 | neuron projection organization | APP,FYN | 0.010 |
| GO:1900449 | regulation of glutamate receptor signaling pathway | APP,FYN | 0.010 |
| HSA-202733 | Cell surface interactions at the vascular wall | FYN,IGHV3-11 | 0.010 |
| GO:0061098 | positive regulation of protein tyrosine kinase activity | APP,FYN | 0.017 |
| GO:1903201 | regulation of oxidative stress-induced cell death | APP,FYN | 0.017 |
| GO:0006897 | Endocytosis | APP,FYN,IGHV3-11 | 0.018 |
| GO:0007631 | feeding behavior | APP,FYN | 0.018 |
| GO:0016358 | dendrite development | APP,FYN | 0.018 |
| GO:0038096 | Fc-gamma receptor signaling pathway involved in phagocytosis | FYN,IGHV3-11 | 0.018 |
| GO:1901216 | positive regulation of neuron death | APP,FYN | 0.018 |
| GO:1903426 | regulation of reactive oxygen species biosynthetic process | APP,FYN | 0.018 |
| GO:1900180 | regulation of protein localization to nucleus | APP,FYN | 0.020 |
| GO:0007612 | Learning | APP,FYN | 0.027 |
| GO:0031347 | regulation of defense response | APP,FYN,IGHV3-11 | 0.027 |
| GO:2001056 | positive regulation of cysteine-type endopeptidase activity | APP,FYN | 0.027 |
| GO:0030162 | regulation of proteolysis | APP,FYN,IGHV3-11 | 0.031 |
| GO:0051897 | positive regulation of protein kinase B signaling | APP,FYN | 0.031 |
| **GO:2000377** | regulation of reactive oxygen species metabolic process | APP,FYN | 0.031 |
| **HSA-168249** | Innate Immune System | APP,FYN,IGHV3-11 | 0.032 |
| **HSA-76002** | Platelet activation, signaling and aggregation | APP,FYN | 0.032 |
| **GO:0050808** | synapse organization | APP,FYN | 0.034 |
| **GO:1901215** | negative regulation of neuron death | APP,FYN | 0.034 |
| **GO:0002684** | positive regulation of immune system process | APP,FYN,IGHV3-11 | 0.037 |
| **GO:0050776** | regulation of immune response | APP,FYN,IGHV3-11 | 0.037 |
| **GO:0002252** | immune effector process | APP,FYN,IGHV3-11 | 0.041 |
| **GO:0007411** | axon guidance | APP,FYN | 0.041 |
| **GO:0019538** | protein metabolic process | APP,BACE1,BACE2,FYN,IGHV3-11 | 0.043 |
| **GO:0006959** | humoral immune response | APP,IGHV3-11 | 0.045 |

**Table 2:** Description of the study population. Information about *TTR* amyloidogenic mutations, sex, age, epigenetically-determined smoking status (never smoker, NS; former smoker, FS; current smoker, CS), and epigenetically-estimated ranges of T cells (CD8T and CD4T), Natural Killer cells (NK), B cells, monocytes (Mono) and granulocytes (Gran) are reported.

| TTR<br>Mutation | N | Sex<br>Female (%) | Age Median<br>(Min-Max) | Smoking |  |  | CD8T Median<br>(Min –Max) | CD4T Median<br>(Min –Max) | NK Median<br>(Min –Max) | B cell Median<br>(Min –Max) | Mono Median<br>(Min –Max) | Gran Median<br>(Min –Max) |
| --- | --- | --- | --- | --- | --- | --- | --- | --- | --- | --- | --- | --- |
|  |  |  |  | NS | FS | CS |  |  |  |  |  |  |
| Cases |  |  |  |  |  |  |  |  |  |  |  |  |
| Val30Met | 33 | 10 (30) | 65<br>(31-88) | 1 | 23 | 9 | 0.033<br>(-3.24×10<sup>-20</sup> – 0.131) | 0.118<br>(1.1×10<sup>-2</sup> – 0.21) | 0.033<br>(-3.81×10<sup>-19</sup> – 0.127) | 1.52×10<sup>-3</sup><br>(-5.55×10<sup>-19</sup> – 0.066) | 0.082<br>(0.038 – 0.147) | 0.634<br>(0.551 – 0.824) |
| Phe64Leu | 8 | 1 (12) | 70<br>(48-75) | 0 | 4 | 4 | 4.02×10<sup>-3</sup><br>(1.07×10<sup>-18</sup> – 1.112) | 0.127<br>(6.9×10<sup>-2</sup> – 0.228) | 0.014<br>(1.96×10<sup>-20</sup> – 0.099) | 3.13×10<sup>-3</sup><br>(-2.17×10<sup>-19</sup> – 0.021) | 0.085<br>(0.057 – 0.107) | 0.65<br>(0.46 – 0.704) |
| Ala120Ser | 2 | 2 (100) | 67-68 | 0 | 2 | 0 | -3.34×10<sup>-20</sup> – 0 | 0.068 – 0.11 | 8.1×10<sup>-3</sup> – 1.28×10<sup>-1</sup> | 0 – 5.1×10<sup>-3</sup> | 0.092 – 0.11 | 0.56 – 0.74 |
| Ile68Leu | 3 | 0 | 53<br>(30-62) | 0 | 0 | 3 | 0 | 0.142<br>(0.07 – 0.16) | 0.038<br>(0.013 – 0.041) | 4.4×10<sup>-3</sup><br>(3.8×10<sup>-3</sup> – 5.1×10<sup>-3</sup>) | 0.083<br>(0.096 – 0.147) | 0.59<br>(0.603 – 0.73) |
| Val122Ile | 1 | 0 | 80 | 0 | 0 | 1 | 0 | 0.23 | 0.042 | 0.005 | 0.071 | 0.55 |
| rs36204272* | 1 | 0 | 65 | 0 | 1 | 0 | 0.069 | 0.14 | 0.052 | 0.006 | 0.129 | 0.505 |
| Controls |  |  |  |  |  |  |  |  |  |  |  |  |
| – | 32 | 19 (59) | 37 (20-76) | 0 | 27 | 5 | 0.015<br>(8.45×10<sup>-19</sup> – 0.13) | 0.16<br>(0.045 – 0.34) | 0.044<br>(-2.68×10<sup>-19</sup> – 0.18) | 0.014<br>(1.08×10<sup>-19</sup> – 0.091) | 0.092<br>(0.051 – 0.13) | 0.571<br>(0.41 – 0.76) |

\*non-coding variant with putative clinical impact

**Figure 1:**
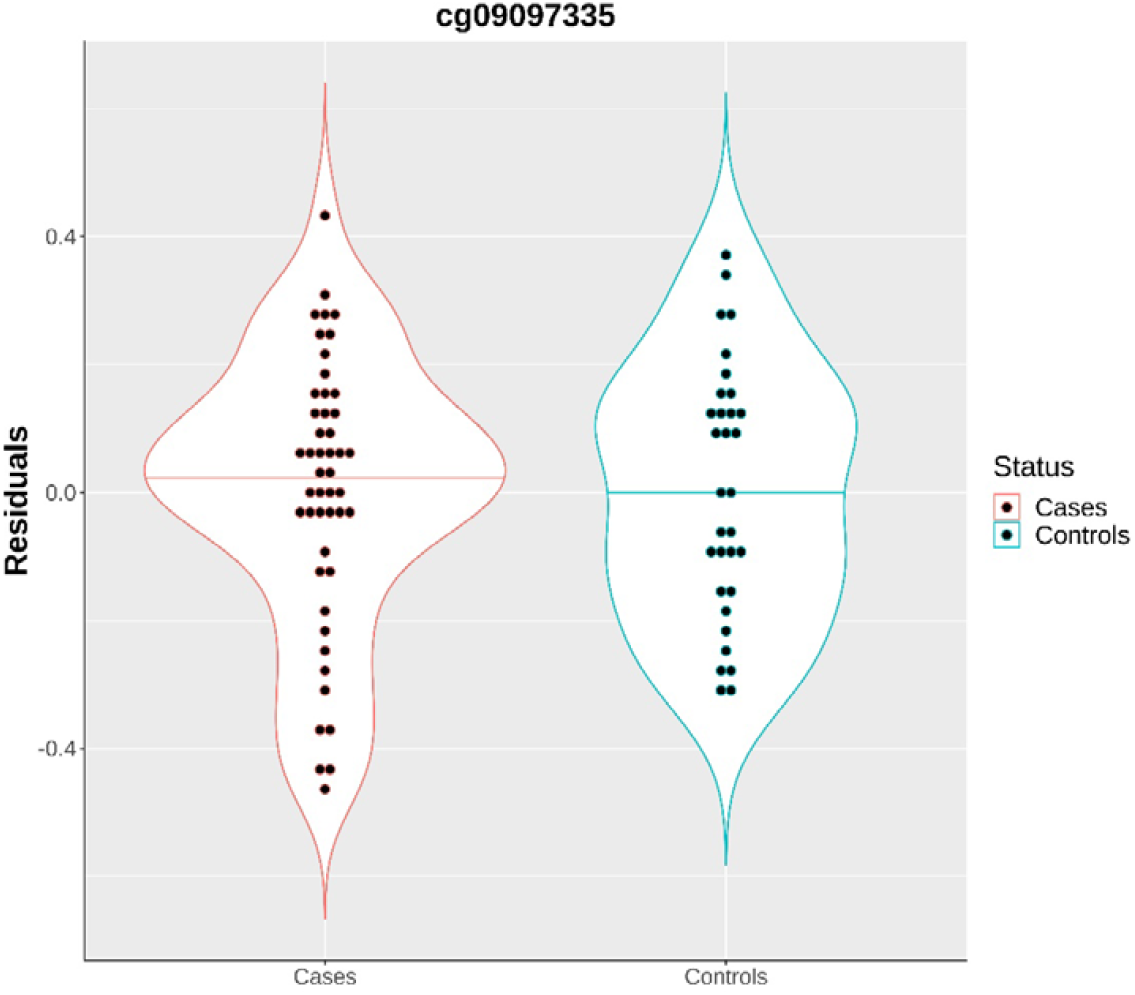
Methylation levels of cg09097335 site in carriers (cases) vs. non-carriers (controls) of amyloidogenic mutations. The residuals were obtained adjusting M values for age, sex, cell composition proportions, top three genetic principal components, and epigenetically-determined smoking status.

**Figure 2:**
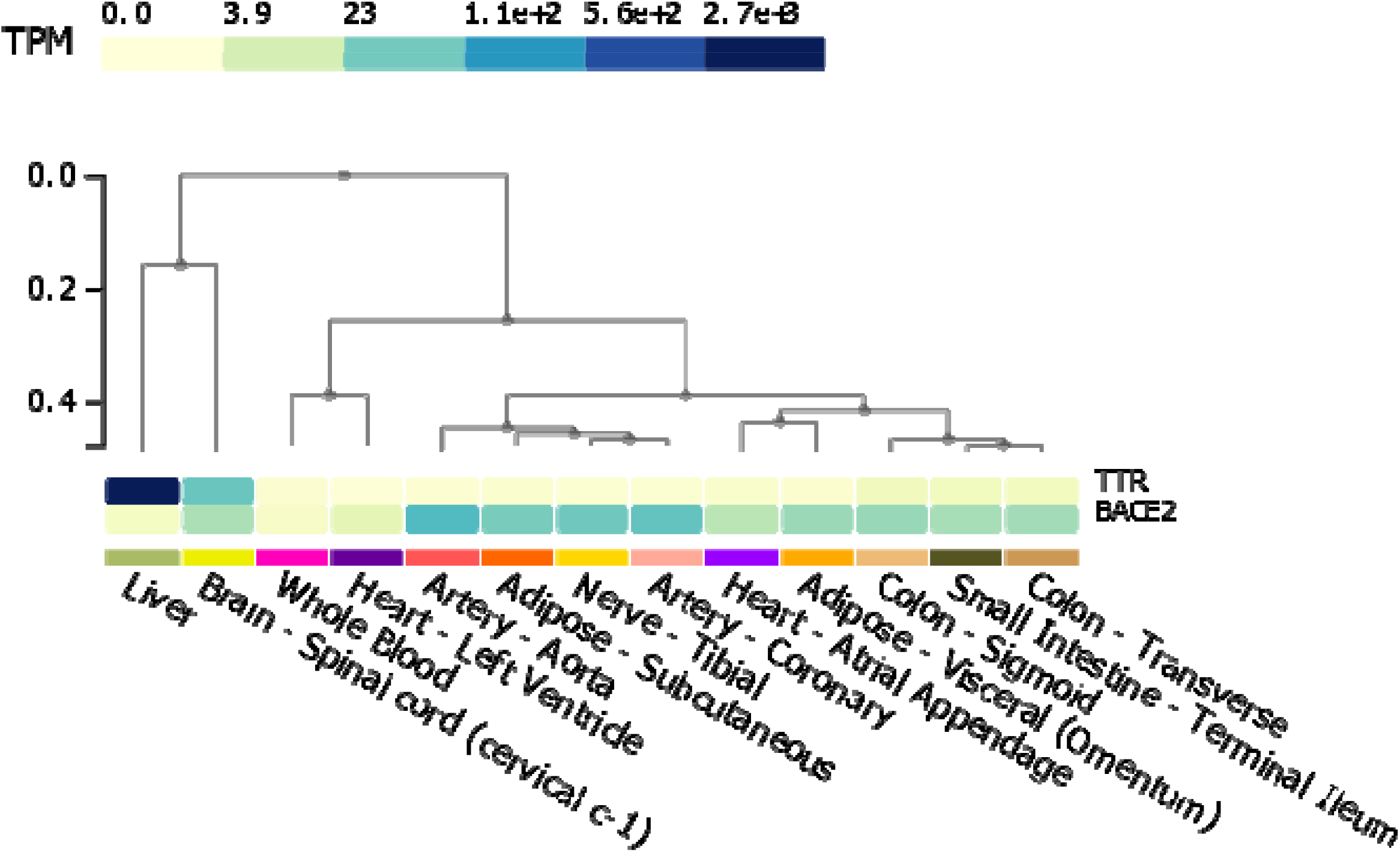
Co-expression of *TTR* and *BACE2* in liver and in hATTR target organs (transcriptomic data from GTEx project, available at https://www.gtexportal.org/).

**Figure 3.**
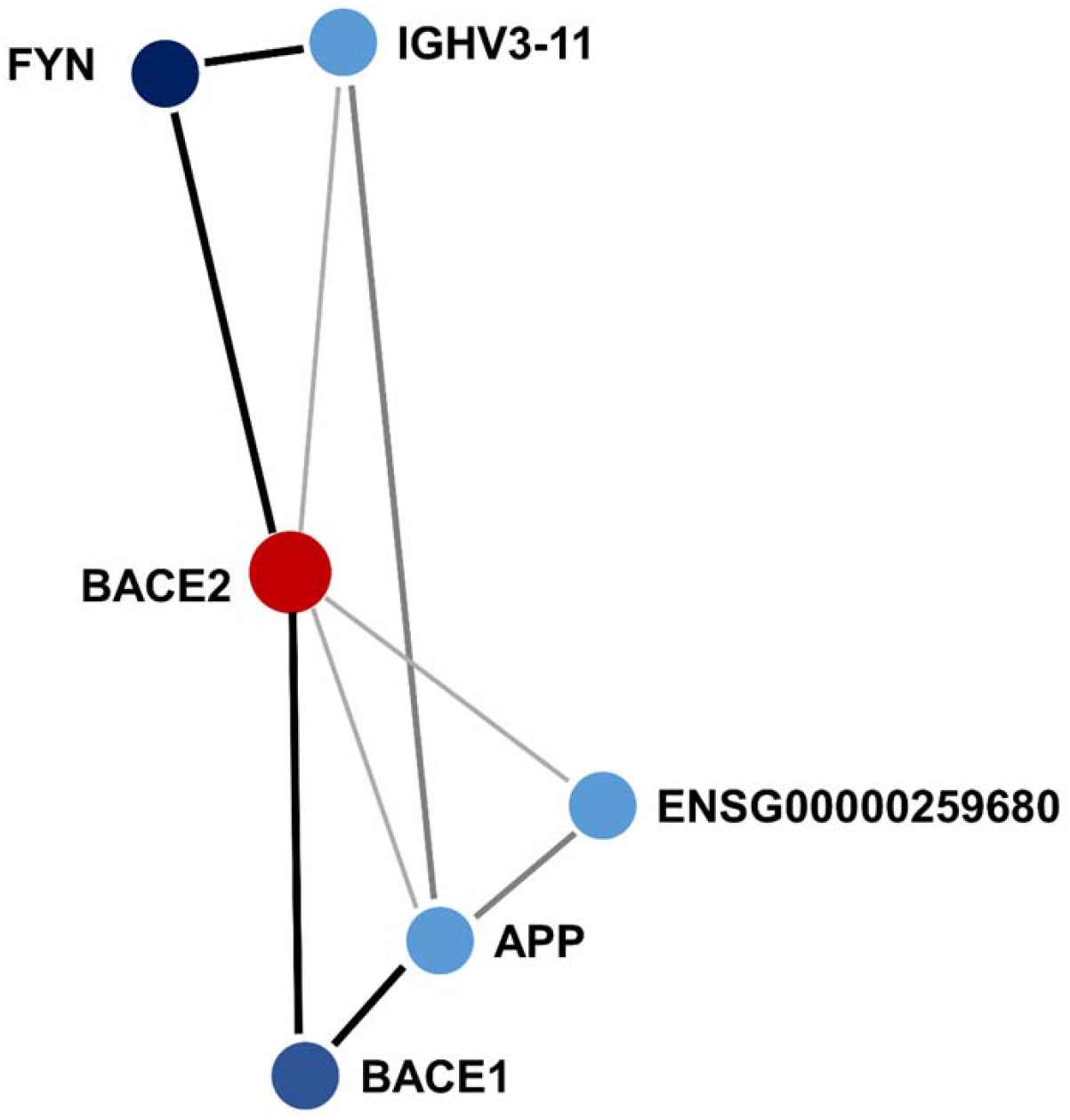
BACE2 protein interaction network. Node colour of the protein is proportional to the interaction score with BACE2. Connector shade and width are proportional to the interaction confidence, highest, high, and medium.

Within *TTR* gene region, we observed that Val30Met mutation disrupts a methylation site, cg13139646, causing a drastic hypomethylation in Val30Met carriers when compared with carriers of other *TTR* mutations (beta= -2.18, p=3.34×10^−11^). To understand whether the disruption of cg13139646 may have functional implications related to hATTR pathogenesis, we performed a co-methylation analysis with respect to cg13139646. We identified 34 methylation sites that are correlated with cg13139646 site (r^2^ > 0.20; Additional File 1). Considering these co-methylated CpG sites, we investigated epigenetic differences among hATTR patients, asymptomatic carriers of *TTR* mutations, and controls (Additional File 2). Applying a Bonferroni correction accounting for the number of CpG sites tested, we observed a significant methylation difference between hATTR patients and asymptomatic carriers of *TTR* amyloidogenic mutations at cg19203115 (r^2^=0.317, beta=-0.555, p=8.6×10^−4^). Nominally significant methylation differences were observed i) between carriers vs. non-carriers at cg11481443 (r^2^=0.263, beta=-0.306, p=3.4×10^−3^) and cg02936398 (r^2^=0.256, beta=0.177, p=4.9×10^−2^); ii) hATTR patients and asymptomatic carriers at cg14311811 (r^2^=0.261, beta=-0.273, p=3.8×10^−2^). Considering symptoms reported by hATTR patients, we identified CpG sites co-methylated with cg13139646 (i.e., the site disrupted by Val30Met mutation) nominally associated with cardiac involvement (cg27392998, r^2^=0.444, beta=-0.235, p=7.5×10^−3^; cg18038361, r^2^=0.433, beta=0.227, p=5×10^−2^), carpal tunnel syndrome (cg16492377; r^2^=0.347, beta=-0.229, p=1.8×10^−2^), and peripheral nervous system involvement (cg14719951, r^2^=0.268, beta=0.249, p=3.5×10^−2^). Since some of these CpG sites were mapped to loci located near *TTR* gene (Additional File 2), we analyzed *TTR* transcriptomic profile in hATTR target organs, observing a different pattern when compared to the expression of the surrounding genes (Additional File 3).

## Discussion

hATTR is a rare multi-organ disorder caused by TTR misfolding and consequently amyloid deposition in several tissues (30). This life-threatening condition is characterized by high clinical heterogeneity with respect to age of onset, penetrance, and phenotypic manifestation (1-10, 30). Although *TTR* amyloidogenic mutations are the cause of *TTR* misfolding, non-coding variation and modifier genes are hypothesized to be involved in wide variability of phenotypic manifestations observed in carriers of *TTR* disease-causing mutations (12, 15, 17-22). Epigenetic modifications (e.g., DNA methylation changes) could also play an important role in the molecular network regulating the hATTR amyloidogenic process (25). To explore this hypothesis, we conducted an EWAS investigating more than 700,000 methylation sites in 48 carriers of *TTR* amyloidogenic mutations and 32 non-carriers. A CpG site (cg09097335) located in *BACE2* gene was significantly hypomethylated in carriers when compared to non-carriers. This gene encodes Beta-secretase 2, a protein mainly known for its role in cleaving APP protein in amyloid-beta, which is a key factor involved in AD pathogenesis (31-33). Several studies have shown that, unlike BACE1 which serves as the primary β*-secretase* protein cleaving APP to amyloid-beta, BACE2 is poorly expressed in the brain and its cleaving ability increases following an inflammatory response (34). APP processing occurs via three proteolytic cleavages caused by α-β- and γ-secretase (35). In non-amyloidogenic processes, α- and γ-secretases lead to the production of a smaller P3 fragment and APP intracellular domain, while, in the amyloidogenic pathway, β-secretase and γ-secretase produce amyloid-beta (35-39). Our results also showed a high-confidence interaction between APP and TTR. Numerous studies explored the interactions between these two amyloidogenic proteins, displaying a relevant biological role of TTR in amyloid-beta aggregation and clearance in AD patients (40-44). Specifically, TTR instability reduces the clearance of amyloid-beta, increasing amyloid toxicity in the brain (40-42). Metal ions and interaction with other proteins could also affect TTR stability (40). Interestingly, a significant association between amyloid-beta levels and AD was identified in AD patients with *TTR* Val30Met (40, 44). A putative amyloidogenic role of amyloid-beta in hATTR was also identified in a post-mortem analysis of a Val30Met carrier where both TTR and amyloid-beta were deposited in the cerebral leptomeningeal and cortical blood vessel walls with a part of the vessel wall occupied by a combination of TTR and amyloid-beta aberrant proteins (43). These previous findings strongly indicate an interplay between the pathogenic mechanisms involved in hATTR and AD. Our epigenome-wide study identified *BACE2* as a potential key factor in this interaction. As previously discussed, BACE2 protein plays a minor role in APP cleaving in the brain (33, 34), while its activity increases in peripheral tissues under inflammatory response (34). Our transcriptomic analysis showed that *BACE2* is expressed in tissues affected by TTR amyloid deposits (i.e., heart, nerves, colon, small intestine, and adipose tissues). Accordingly, the methylation change observed in the TTR-mutation carriers is possibly due to the role of BACE2 in response to the inflammation induced by TTR amyloidogenic process in peripheral tissues (45).

Within *TTR* gene region, we observed that Val30Met disrupts a CpG site, causing a drastic hypomethylation in the carriers of this mutation. SNPs at CpG sites disrupting the methylation reactions can be associated with changes in regulatory function. (46, 47). To explore the functional consequences of cg13139646 disruption, we analyzed the co-methylation of this site with CpG sites in the surrounding regions (NC_000018.9: 28,171,000–30,171,500). Indeed, co-methylation patterns reflect specific molecular mechanisms responsible for the regulation of multiple genes located in the same region (48). In our analysis, some of the CpG sites identified map to *TTR* gene region, while others map in nearby loci. Considering hATTR target organs, these surrounding loci with co-methylated CpG sites have higher gene expression than *TTR*. We speculate that these genes may be involved in the formation of TTR amyloid deposits. This hypothesis is supported by the fact that co-methylated CpG sites are associated with hATTR traits. The strongest evidence was observed with respect to cg19203115 mapped in *B4GALT6* gene. Considering a Bonferroni correction accounting for the number of co-methylated CpG sites tested, cg19203115 showed a significant difference in methylation levels between hATTR patients and asymptomatic carriers of *TTR* mutations. *B4GALT6* gene encodes Beta-1,4-Galactosyltransferase 6, a type II membrane-bound glycoprotein that has exclusive specificity for the donor substrate UDP-galactose. The enzyme activity of B4GALT6 changes in response to inflammatory processes (49). B4GALT6 stimulates astrocyte activation through the catalyzation of lactosylceramide synthesis, which in turn controls the production of pro-inflammatory cytokines and chemokines (49). Hence, observing methylation changes in B4GALT6 may be associated with inflammatory response to TTR amyloid deposits. Nominally significant differences were observed for CpG sites mapped in other surrounding loci: *DSC2* (cg02936398, Carriers *vs*. Controls); *DSG2* (cg14311811, hATTR patients *vs*. asymptomatic carriers); *DSC3* (cg16492377, carpal tunnel syndrome in hATTR patients). *DSC2* and *DSG2* encode components of the desmosome. This protein complex is specialized for cell-to-cell adhesion in myocardial tissue and mutations in *DSC2* and *DSG2* genes are associated with arrhythmogenic right ventricular cardiomyopathy (50). In an *in vivo* study of myocardial inflammation, *DSC2* overexpression was observed to lead to tissue necrosis, fibrosis, and calcification of ventricles (51). This process alters homeostasis among desmosomal proteins, inducing a cascade of different cell-cell interactions leading to cardiac remodeling (51). In hATTR, cardiac amyloid fibril depositions also led to tissue dysfunctions (7). Heart failure, restrictive cardiomyopathy, and rhythm disturbances (i.e. conduction system diseases, atrial fibrillation, and ventricular tachycardia) are the main clinical signs that occur after the accumulation of misfolded TTR protein (52-54). Furthermore, transcriptomic interaction is observed between TTR and DSG2 to induce hypertrophic cardiomyopathy in animal models (55). In this context, methylation changes in *DSC2* and *DSG2* genes could reflect pathogenic processes in hATTR target organs. We also identified two CpG sites co-methylated with cg13139646 (i.e., the methylation site disrupted by Val30Met mutation) that are nominally associated with hATTR symptoms. Cg18038361 is located in *TTR* gene promoter region and is associated with cardiac involvement in hATTR patients. DNA methylation changes in promoter regions are well-known to play an important role in gene expression regulation (56, 57). Cg18038361 association may be linked to regulatory changes in *TTR* gene expression. Lastly, two CpG sites - cg16492377 and cg14719951, map to *DSC3* transcription start site and gene body, respectively. In our analysis, methylation changes in these sites were associated with carpal tunnel syndrome and peripheral nervous system involvement in hATTR patients, respectively. *DSC3* gene encodes the desmocollin-3 a calcium-dependent glycoprotein. Low *DSC3* expression in human epidermis leads to a loss of tissue integrity (58). We speculate that cg16492377 methylation association with carpal tunnel syndrome may be related to changes in *DSC3* transcriptomic regulation. Although we provide novel findings regarding the role of methylation changes in hATTR, our study presents several limitations. Since hATTR is a rare disease, we investigated a relatively small sample size. Our calculation showed that the sample size investigated in our main analysis should provide >80% statistical power to detect medium effect sizes (Δ_β_=0.2; Additional File 4). However, large samples will be needed to investigate how epigenetic changes affect hATTR symptoms and differences across *TTR* amyloidogenic mutations. Our cohort showed age and sex differences between carriers of *TTR* amyloidogenic mutations and controls. Our analysis was adjusted for these confounding variables together with blood cell types, genetic principal components, and epigenetically-determined smoking status. More balanced case-control groups are needed to investigate how epigenetic differences are associated differently between sexes and across age groups. We used transcriptomic data from GTEx project to explore the potential mechanisms related to the epigenetic associations identified. Further studies generating transcriptomic and epigenomic information across multiple informative tissues will provide a more comprehensive understanding of the molecular processes involved in hATTR. The CpG site disrupted by Val30Met mutation (i.e., cg13139646) is co-methylated with CpG sites associated with hATTR and its symptoms. We speculated possible regulatory mechanisms, also discussing the potential role of *TTR*-surrounding genes as hATTR modifier loci. These findings should be considered preliminary results that need to be replicated and expanded by future investigations.

## Conclusions

Our study provided novel insights regarding the pathogenesis of hATTR, supporting the involvement of methylation changes in the amyloidogenic process induced by *TTR* disease-causing mutations. Further studies will be needed to characterize specific mechanisms underlying the epigenetic associations, in particular, the potential role of amyloid-beta metabolic process and inflammatory response. The understanding of how methylation changes modulate the penetrance and the severity of *TTR* mutations could lead to the identification of novel targets to develop treatments and screening tools for the carriers.

## Methods

Thirty-eight symptomatic patients and 10 asymptomatic *TTR* mutations carriers were recruited from three Italian centers for the treatment of systemic amyloidosis: “San Giovanni Calibita” Fatebenefratelli Hospital, Isola Tiberina – Rome, Fondazione Policlinico Universitario “A. Gemelli” – Rome and Careggi University Hospital – Florence (16-20). Thirty-two controls were recruited by the Department of Biology – University of Rome “Tor Vergata” (Table 3). hATTR diagnosis was based on the presence of clinical signs and symptoms and the presence of an amyloidogenic mutation on *TTR* gene. The coding mutations identified include: Val30Met (rs28933979), Phe64Leu (rs121918091), Ile68Leu (rs121918085), Ala120Ser (rs876658108), and Val122Ile (rs76992529). One hATTR patient is a carrier of a mutation (rs36204272) in an intronic region with a putative clinical impact (59). Carpal tunnel syndrome and cardiac involvement with confirmed TTR amyloid deposits are present in rs36204272 carrier. Information regarding the organ involvements was collected for each patient: peripheral and nerve involvement (nerve conduction study); cardiac involvement (electrocardiographic and echocardiography anomalies); gastrointestinal involvement (gastric paresis, stypsis, or diarrhea); autonomic neurological involvement (orthostatic hypotension and urinary incontinence); ocular involvement (vitreous opacities): and carpal tunnel syndrome (median nerve decompression) (11, 60-62). The present study was approved under the protocol 39/18 by the Comitato Etico Indipendente, Fondazione Policlinico Tor Vergata – Rome, Italy.

### DNA methylation analysis

DNA was extracted using the phenol/chloroform protocol (63) and purified through Amicon Ultra-0.5 mL Centrifugal Filters (EMD Millipore) to achieve a DNA concentration of 100 ng/µL. DNA concentration was checked via NanoDrop technology (ND-1000, Thermofisher Scientific) and Qubit Quantitation technology (High Accuracy & Sensitivity, Thermofisher). DNA methylation analysis was executed in two phases: the EZ DNA Methylation kit (Zymo Research) was used to perform sodium bisulfite conversion; the Illumina Infinium Methylation EPIC Chip (with over 850,000 methylation sites; Illumina Inc.) was used to quantify DNA methylation according to the standard Illumina protocol. The methylation array analysis was performed at the Connecting bio-research and Industry Center, Trieste – Italy.

### Preprocessing, quality control, and normalization

The raw signal intensity files were processed and cleaned using R 3.6 with ChAMP package (64). The ratio of methylated and unmethylated intensities from idat files was converted into beta values for further processing. The probes failing thresholds on detection value, bead count, sites near SNPs, probes that align to multiple positions, sex chromosomes and outliers were removed. None of the samples failed quality control. The remaining 718,509 probes for 80 individuals were normalized with BMIQ. Batch effects were assessed using singular vector decomposition and corrected with ComBat method (65). The genomic lambda of the case-control association was 1.03, calculated using QQPerm package (https://cran.r-project.org/web/packages/QQperm/index.html).

### Blood cell type composition, genetic variability estimation, and smoking prediction

References-based method was employed to adjust for the heterogeneity due to the cell type composition of the whole blood samples investigated (66). This method uses specific DNA methylation signatures derived from purified whole blood cell-type as biomarkers of cell identity, to correct beta value dataset. Cell proportions for five cell-types (B cells, granulocytes, monocytes, natural killer cells, and T cells) were detected, and a linear regression was applied (64, 66). To account for the genetic variability among the samples investigated, principal components (PCs) were calculate using the method proposed by Barfield, Almli (67). This approach allowed us to compute PCs based on CpGs selected for their proximity to SNPs. The data obtained can be used to adjust for population stratification in DNA methylation studies when genome-wide SNP data are unavailable (67). Cigarette smoke has a very large effect on DNA methylation profile, triggering alteration at multiple CpGs (68). Consequently, smoking status needs to be considered as a potential confounder in epigenetic association studies. EpiSmokEr package was used to classify the smoking status of each participant on the basis of their epigenetic profile (69). Briefly, EpiSmoker is a prediction tool that provides smoking probabilities for each individual (never-smoker, former-smoker, and current smoker) using a set of 121 informative CpG sites (68).

### Data analysis

We conducted two epigenome-wide analyses testing 718,509 methylation sites. First, we investigated the methylation changes between 48 carriers of a *TTR* amyloidogenic mutation) and 32 controls (i.e., non-carriers). Considering CpG sites that survive epigenome-wide multiple testing correction, we also verified whether the associations observed were due to disease-associated genetic differences or post-disease processes, comparing i) patients affected by hATTR *vs*. controls, ii) asymptomatic carriers of *TTR* mutations *vs*. controls, and iii) patients affected by hATTR *vs*. asymptomatic carriers of *TTR* mutations. To investigate the functionality of the CpG disrupted by Val30Met mutation, we analyzed its co-methylation with CpG sites in the surrounding region (NC_000018.9: 28,171,000–30,171,500). This region was selected based on the *TTR* regulatory mechanisms observed in previous studies (20-22). We used *cor()* R function to calculate Pearson’s correlation coefficient testing 367 sites and considering the methylation levels (M values) in the controls. CpG sites with high co-methylation (r^2^>0.2) were investigated with respect to hATTR-related traits (i.e., carrier status, disease status, and symptoms). In all association analyses, we implemented a linear regression analysis including cell composition proportions, top three genetic PCs, epigenetically-determined smoking status, age, and sex as covariates. FDR method (70) was applied to adjust the results for epigenome-wide testing and the q-value < 0.05 was considered as the significance threshold. Co-expression analysis was conducted using GTEx v8 (29) via the Multi Gene Query available at https://www.gtexportal.org/. *Ggplot2* R package (71) was employed to plot co-methylation pattern results. STRING v.11.0 (72) was used to identify protein interaction with the loci identified, considering experiments, co-expression, co-occurrence, gene fusion, and neighborhood as active sources and an interaction score higher than 0.4 (medium confidence). The protein interaction network was investigated further conducting functional enrichments association related to the protein-protein interactions identified considering Gene Ontologies (73) for biological processes and molecular pathways available from Reactome Database (74) and Kyoto Encyclopedia of Genes and Genomes (KEGG) (75). FDR (q-value < 0.05) was applied to account for multiple testing assuming the whole genome as the statistical background. Statistical power calculation was done using pwrEWAS tool (76) considering medium and small effect sizes (Δ_β_ = 0.5 and 0.2, respectively) and multiple sample sizes.

## Data Availability

Data supporting the findings of this study are available within this article and its additional files.

## List of abbreviations

AD: Alzheimer disease
APP: amyloid beta precursor protein
hATTR: Hereditary transthyretin amyloidosis
BACE1: beta-secretase 1
B4GALT6: beta-1,4-Galactosyltransferase 6
BACE2: beta-secretase 2
DSC2: descmocollin-2
DSC3: desmocollin-3
DSG3: desmoglein-3
EWAS: epigenome-wide association studies
FDR: false discovery rate
FYN: FYN proto-oncogene, Src family tyrosine kinase
IGHV3-11: immunoglobulin heavy variable 3-11
TTR: transthyretin

## Declarations

### Ethics approval and consent to participate

This study was approved under the protocol 39/18 by the Comitato Etico Indipendente, Fondazione Policlinico Tor Vergata – Rome, Italy. Informed consent was obtained from each participant involved.

### Authors’ contributions

ADL, FDA, MF, and RP were involved in study design. MDG, ML, MS, FP, SF, and DM conducted the recruitment and assessment of the participants. ADL, GAP, and RP carried out the statistical analysis. All authors were involved in the interpretation of the results. ADL and RP wrote the first draft of the manuscript and all authors contributed to the final version of the manuscript.

## Acknowledgements

We thank the participants involved in this study and their caregivers.

